# Brain morphometry is not associated with non-motor outcomes after deep brain stimulation in Parkinson’s disease

**DOI:** 10.1101/2023.06.07.23291019

**Authors:** Philipp Alexander Loehrer, Wibke Schumacher, Stefanie T. Jost, Monty Silverdale, Jan Niklas Petry-Schmelzer, Anna Sauerbier, Alexandra Gronostay, Veerle Visser-Vandewalle, Gereon R. Fink, Julian Evans, Max Krause, Alexandra Rizos, Angelo Antonini, Keyoumars Ashkan, Pablo Martinez-Martin, Christian Gaser, K. Ray Chaudhuri, Lars Timmermann, Carlos Baldermann, Haidar S. Dafsari, EUROPAR and the International Parkinson and Movement Disorders Society Non-Motor Parkinson’s Disease Study Group

## Abstract

Deep brain stimulation of the subthalamic nucleus (STN-DBS) is an established therapy in advanced Parkinson’s disease (PD). Motor and non-motor outcomes, however, show considerable inter-individual variability. Morphometry-based metrics have recently received increasing attention to predict treatment effects. As evidence for the prediction of non-motor outcomes is limited, we sought to investigate the association between metrics of voxel-based morphometry and short-term non-motor outcomes following STN-DBS in this prospective open-label study. 49 PD patients underwent structural MRI and a comprehensive clinical assessment at preoperative baseline and 6-month follow-up. Voxel-based morphometry was used to assess associations between cerebral volume and non-motor outcomes corrected for multiple comparisons using a permutation-based approach. We replicated existing results associating atrophy of the superior frontal cortex with subpar motor outcomes. Non-motor outcomes, however, were not associated with morphometric features, limiting its use as a marker to inform patient selection and holistic preoperative counselling.

## Introduction

Deep brain stimulation (DBS) of the subthalamic nucleus (STN) is an established therapy for the treatment of motor and non-motor symptoms in advanced Parkinson’s disease (PD).^1–3^ Despite its well-established effects at the group level, individual symptom relief varies significantly, complicating preoperative patient selection and counselling.^4^ To predict outcomes and support preoperative management, neuroimaging-based biomarkers using advanced imaging technologies, such as tractography and functional MRI, have proven useful.^4, 5^ Their widespread clinical application, however, is limited by the need for additional and sometimes time-consuming scanning protocols and the expertise to analyse and translate their results.^6^ Therefore, the association of postoperative outcomes with metrics based on T1-weighted sequences obtained in clinical routine during surgical planning has been investigated. Particularly, analyses focussing on morphometric tissue features, such as voxel-based morphometry (VBM), have lately received increasing attention to monitor clinical progression and treatment effects.^6^ In a recent meta-analysis including 1253 patients enrolled in 24 studies, Wang and colleagues identified specific areas whose morphometric features were associated with outcomes following STN-DBS.^6^ Here, atrophy of the motor cortex and thalamus was associated with below-average improvement in motor symptoms. On the other hand, outcome prediction of non-motor symptoms has received little attention, with studies focusing on cognitive decline and immediate psychiatric alterations such as postoperative confusion, delirium, and impulsivity.^6^ Poor outcomes in verbal memory were associated with hippocampal atrophy at baseline, while immediate psychiatric complications were related to caudal middle frontal cortex atrophy.^6^ As STN-DBS is associated with beneficial short-term outcomes in a range of non-motor symptoms such as sleep/fatigue, attention/memory, and mood/apathy,^7, 8^ in the present study, we sought to explore the association between a large spectrum of non-motor symptoms and volumetric properties.

**Figure 1.**
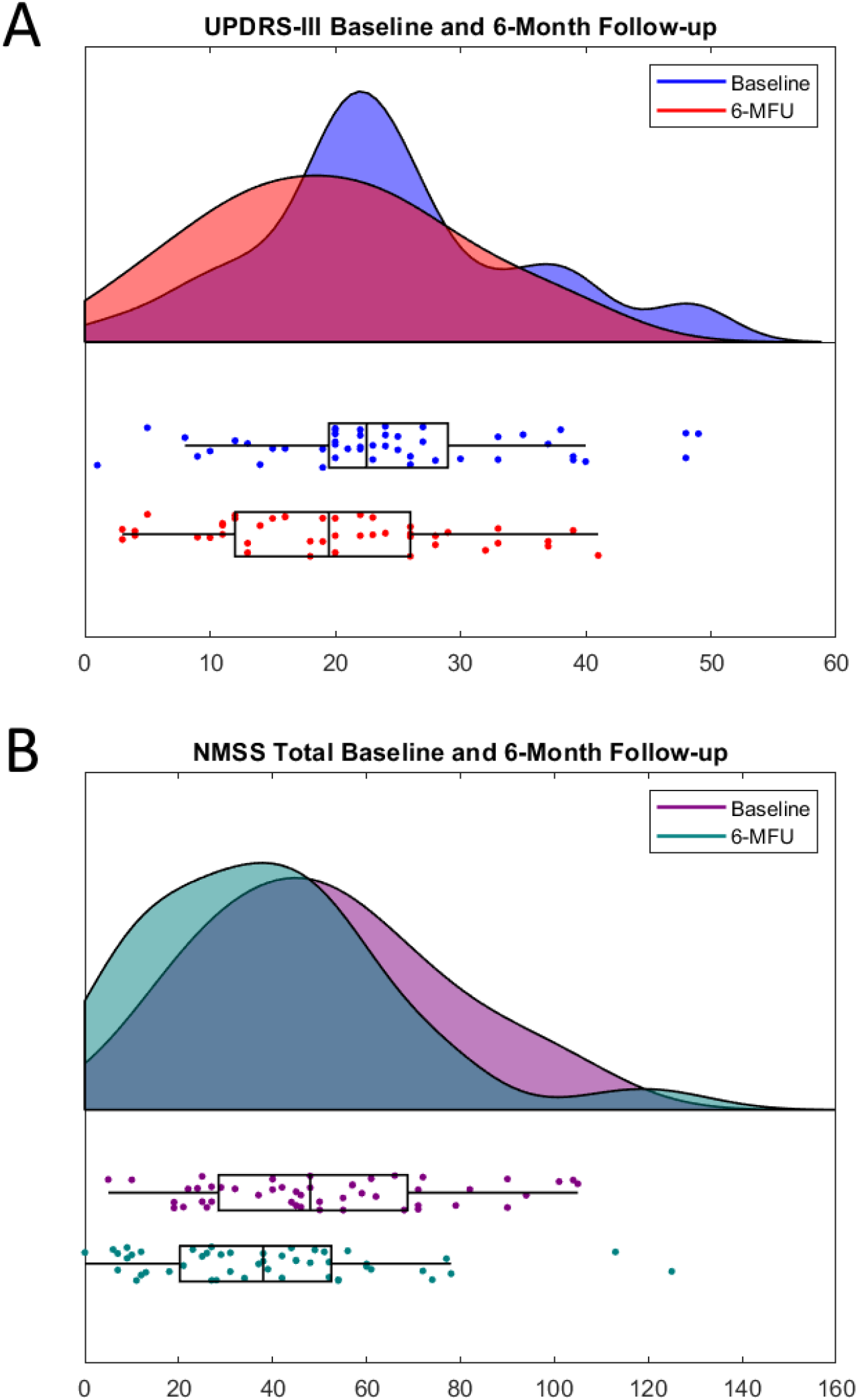
Visualisation of baseline and 6-month follow-up values of the Unified Parkinson’s Disease Rating Scale-part III **(A)** and Non-Motor Symptom Scale-total score **(B)**.

**Figure 2.**
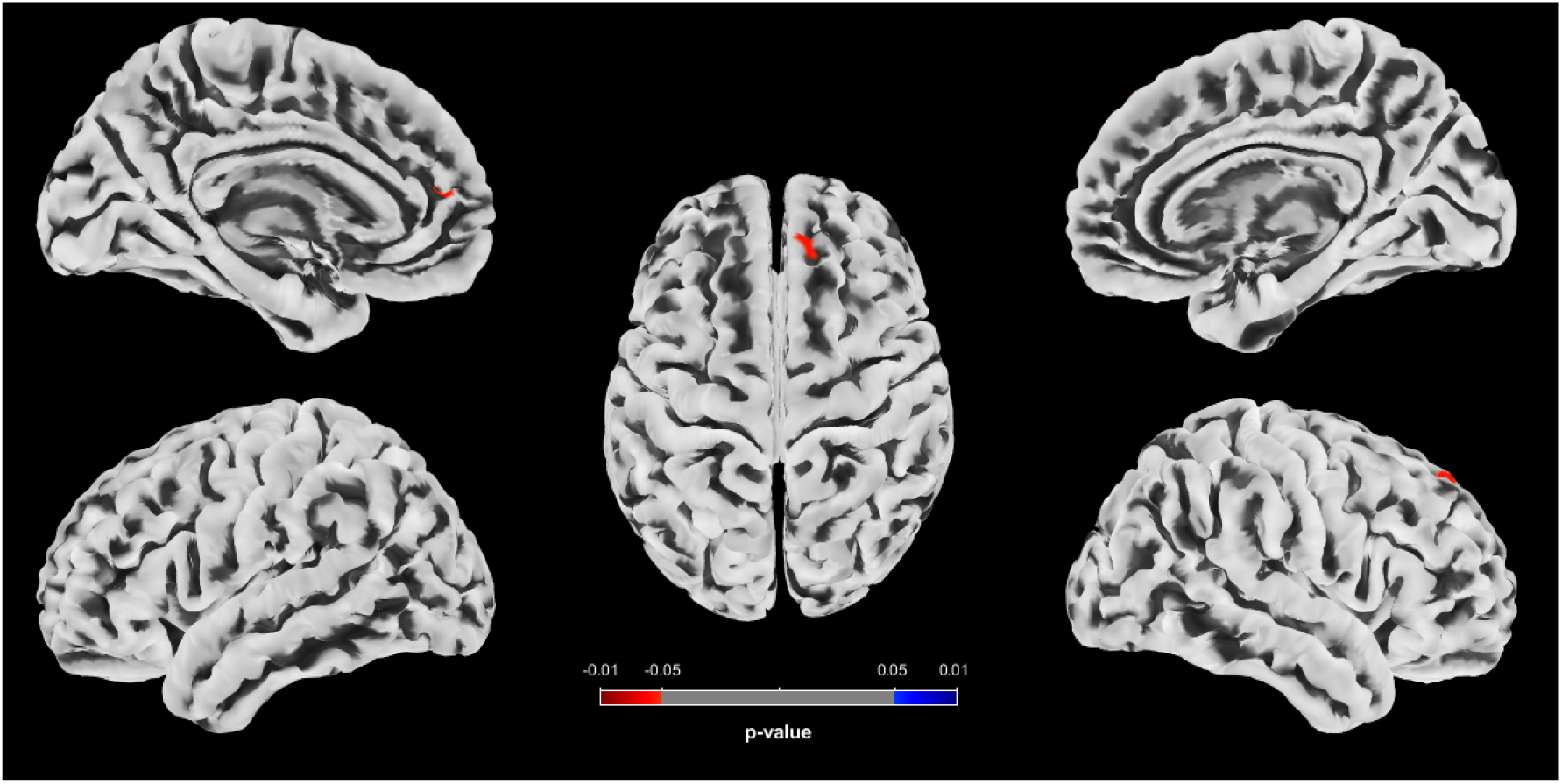
Clusters with an association between metrics of voxel-based morphometry and postoperative change in UPDRS-III as revealed by the whole brain analysis. Results are displayed as surface overlays. Clusters denote regions of grey matter loss significantly associated with poor motor response to deep brain stimulation. P-Values were corrected for multiple comparisons using a permutation-based approach and thresholded at p<.05, family-wise error-corrected.

## Results

### Clinical outcomes

Forty-nine patients with PD (31 males, mean age 64.5 ± 8.2 years) were enrolled. At the six month follow-up, the following scales improved: NMSS total score (p=.006, Cohen’s d=.46), PDQ-8 SI (p<.001, Cohen’s d=.52), UPDRS III (p=.023, Cohen’s d=.45), SCOPA-M activities of daily living (p<.001, Cohen’s d=.56), SCOPA-M motor complications (p<.001, Cohen’s d=.85), and LEDD (p<.001, Cohen’s d=1.06). Analysis of NMSS domains revealed beneficial effects of STN-DBS on sleep/fatigue (p=.003, Cohen’s d=.55), perceptual problems/hallucinations (p=.023, Cohen’s d=.39), urinary symptoms (p=.023, Cohen’s d=.35), and miscellaneous symptoms (p<.001, Cohen’s d=.68). Longitudinal changes of clinical outcomes are reported in Table 1.

**Table 1:**
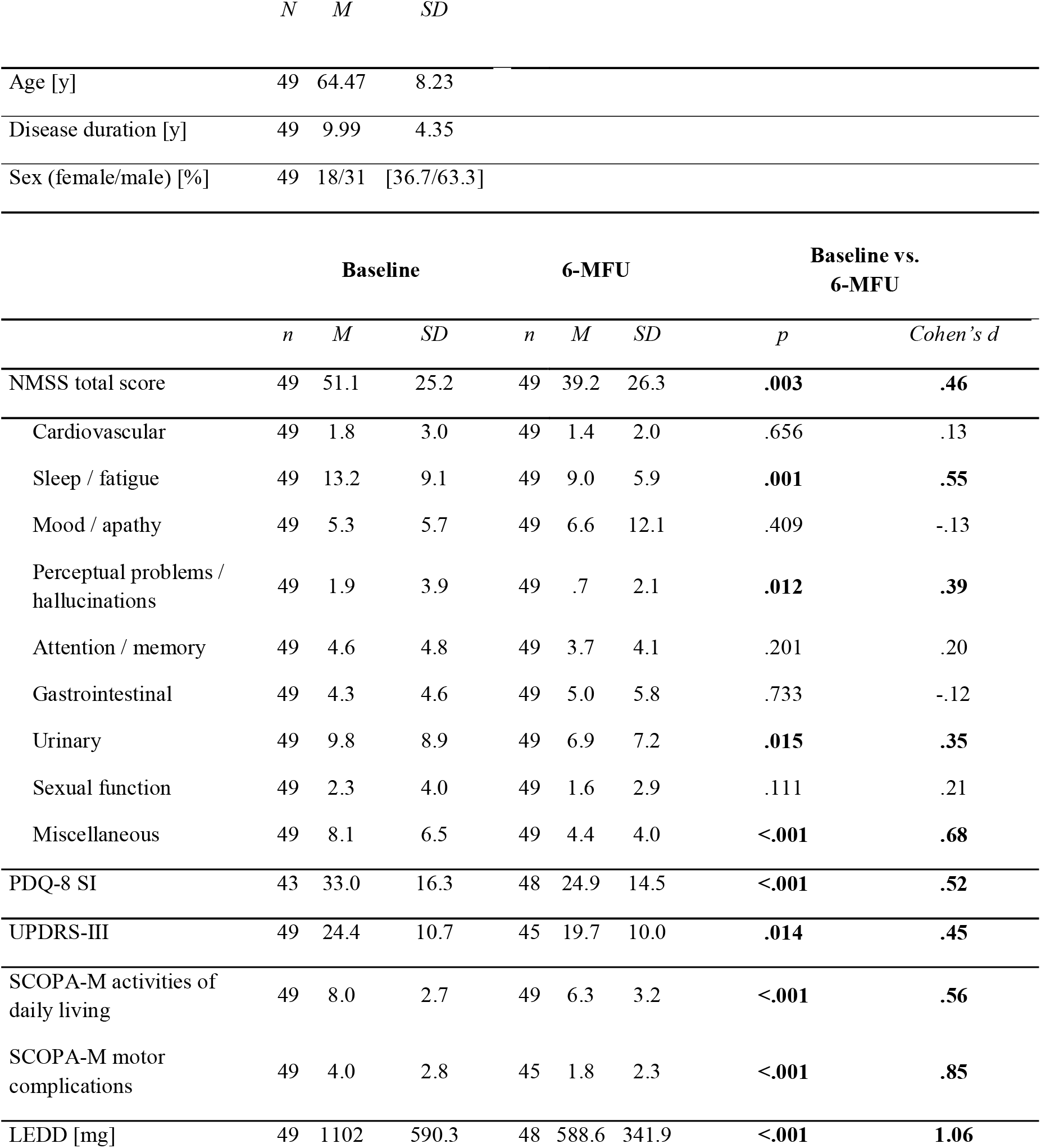
Baseline characteristics and outcomes at baseline and 6-month follow-up. Demographic characteristics and outcome parameters at baseline and 6-month follow-up. Reported p-values are corrected for multiple comparisons using Benjamini-Hochberg’s method (six scales). Bold font highlights significant results, p<.05. **Abbreviations:** 6-MFU = 6-month follow-up; LEDD = Levodopa equivalent daily dose; LEDD-DA: LEDD of dopamine agonists; NMSS = Non-Motor Symptom Scale; PDQ-8 SI = 8-item Parkinson’s Disease Questionnaire Summary Index; SCOPA-M = Scales for Outcomes in Parkinson’s disease-Motor; SD = Standard deviation, UPDRS III = Unified Parkinson’s Disease Rating Scale Part III

|  | *N* | *M* | *SD* |  |  |  |  |  |
| --- | --- | --- | --- | --- | --- | --- | --- | --- |
| Age [y] | 49 | 64.47 | 8.23 |  |  |  |  |  |
| Disease duration [y] | 49 | 9.99 | 4.35 |  |  |  |  |  |
| Sex (female/male) [%] | 49 | 18/31 | [36.7/63.3] |  |  |  |  |  |
|  | Baseline |  |  | 6-MFU |  |  | Baseline vs. 6-MFU |  |
|  | *n* | *M* | *SD* | *n* | *M* | *SD* | *p* | *Cohen's d* |
| NMSS total score | 49 | 51.1 | 25.2 | 49 | 39.2 | 26.3 | **.003** | **.46** |
| Cardiovascular | 49 | 1.8 | 3.0 | 49 | 1.4 | 2.0 | .656 | .13 |
| Sleep / fatigue | 49 | 13.2 | 9.1 | 49 | 9.0 | 5.9 | **.001** | **.55** |
| Mood / apathy | 49 | 5.3 | 5.7 | 49 | 6.6 | 12.1 | .409 | -.13 |
| Perceptual problems / hallucinations | 49 | 1.9 | 3.9 | 49 | .7 | 2.1 | **.012** | **.39** |
| Attention / memory | 49 | 4.6 | 4.8 | 49 | 3.7 | 4.1 | .201 | .20 |
| Gastrointestinal | 49 | 4.3 | 4.6 | 49 | 5.0 | 5.8 | .733 | -.12 |
| Urinary | 49 | 9.8 | 8.9 | 49 | 6.9 | 7.2 | **.015** | **.35** |
| Sexual function | 49 | 2.3 | 4.0 | 49 | 1.6 | 2.9 | .111 | .21 |
| Miscellaneous | 49 | 8.1 | 6.5 | 49 | 4.4 | 4.0 | **<.001** | **.68** |
| PDQ-8 SI | 43 | 33.0 | 16.3 | 48 | 24.9 | 14.5 | **<.001** | **.52** |
| UPDRS-III | 49 | 24.4 | 10.7 | 45 | 19.7 | 10.0 | **.014** | **.45** |
| SCOPA-M activities of daily living | 49 | 8.0 | 2.7 | 49 | 6.3 | 3.2 | **<.001** | **.56** |
| SCOPA-M motor complications | 49 | 4.0 | 2.8 | 45 | 1.8 | 2.3 | **<.001** | **.85** |
| LEDD [mg] | 49 | 1102 | 590.3 | 48 | 588.6 | 341.9 | **<.001** | **1.06** |

**Table 2:** Association between brain morphometry and UPDRS-III. Characteristics of clusters with an association between metrics of voxel-based morphometry and postoperative change in UPDRS-III. “Cluster” denotes clusters with a significant association between grey matter loss and poor motor response to deep brain stimulation (DBS). “Location” indicates the anatomical landmark comprising the majority of voxels of a cluster, according to the Desikan-Killiany Atlas. P-Values are clusterwise p-values corrected for multiple comparisons. “MNI152-coordinates” describe the coordinates of the cluster’s center of gravity in MNI152-space.

| Cluster | Location | p-Value | MNI152-Coordinates |  |  |
| --- | --- | --- | --- | --- | --- |
|  |  |  | X | Y | Z |
| 1 | Left superior frontal cortex | .029 | -14 | 51 | 17 |
| 2 | Right superior frontal cortex | .036 | 12 | 41 | 48 |

### Association of morphometry surrogates and postoperative motor symptom change

A multiple regression analysis assessed the relationship between motor response to STN-DBS and morphometry surrogates. Following a threshold-free cluster enhancement (TFCE) approach to correct for multiple comparisons, a cluster within the bilateral superior frontal cortex showed a positive association with changes in postoperative motor function.

### Association of morphometry surrogates and postoperative non-motor symptom change

No significant associations were observed between morphometry surrogates and change scores of NMSS-T and NMSS-domains, following a TFCE approach to correct for multiple comparisons.

## Discussion

In the present study, we investigated the association between brain morphometric features and changes in clinical outcomes following STN-DBS in PD. We replicated findings that impaired integrity of the frontal cortex is associated with subpar improvement in motor symptoms following STN-DBS. In contrast, changes in non-motor symptoms were not associated with features of brain morphometry.

Employing a whole brain voxel-based morphometry analysis, we identified an association between reduced volume of the bilateral superior frontal cortex and poor motor outcomes after neurostimulation in PD. This finding is consistent with previous studies demonstrating an association between poor motor outcomes and reduced cortical thickness and a diminished volume of this region.^9, 10^ Importantly, the superior frontal cortex comprises the critical structures of the motor network for movement generation and control.^9, 11, 12^ Information processing within this network is altered in PD and modulated by dopaminergic replacement therapy and DBS, resulting in improved motor function.^13, 14^ In STN-DBS, this modulation is in part elicited by antidromic conveyance of stimulation signals via the hyperdirect pathway that directly links the subthalamic nucleus to structures of the motor network.^15^ Consequently, integrity of the frontal cortex seems crucial for STN-DBS to exert its effects and might serve as a marker to predict motor response after DBS surgery. In future studies, larger sample sizes in multicentre cohorts are needed to define patient-specific thresholds and thus implement bilateral superior frontal cortex volume as a biomarker for individual outcome prediction.

Contrary to our expectations, there were no associations between morphometric features and postoperative changes in non-motor symptoms. The effect sizes of postoperative motor and non-motor symptom change were similar and we observed the aforementioned association between motor symptoms and brain morphometric features. Therefore, we reason that our findings for non-motor symptoms genuinely reflect the absence of an effect rather than being attributed to low sensitivity. As non-motor symptoms constitute a heterogeneous group of symptoms,^16, 17^ several aspects have to be considered when interpreting the present findings.

First, this study investigated the brain areas associated with postoperative changes in a wide range of non-motor symptoms assessed by the NMSS total and its domain scores. Previous studies found limited evidence of an association between cortical atrophy and postoperative changes in various cognitive functions.^18^ Lower hippocampal volume, however, has been associated with a postoperative decline in verbal memory.^18, 19^ In the present study, lower hippocampal volumes were associated with less postoperative changes in the attention and memory domains, although these results did not survive TFCE correction. The lack of significant results in this domain might be attributed to the presumably low sensitivity of the NMSS to detect subtle changes in a single cognitive domain. The NMSS is a clinician-rated scale investigating a wide range of NMS across nine domains whereby symptoms are rated according to severity and frequency. Thus, it is not intended to evaluate specific cognitive domains but global cognition and to assess the progress or treatment response of a wide range of non-motor symptoms.^20^

Second, the present study investigated the association between brain morphometry and short-term non-motor outcomes, not the development or worsening of pre-existing non-motor symptoms, which may result from progression of Parkinson’s disease rather than from neurostimulation. In this context, Aybek and colleagues identified hippocampal volume as a marker to predict postsurgical conversion to dementia in a long-term, i.e., 25 months, follow-up.^21^ Patients with Parkinson’s disease dementia had smaller preoperative hippocampal volumes than patients without conversion. The authors concluded that hippocampal atrophy is a potential clinical marker to predict postoperative conversion to dementia, but that the postsurgical development is due to the disease progression rather than the procedure itself.^21^ As the present study investigated short-term outcomes only, further studies investigating the relationship between brain morphometry and long-term non-motor outcomes are needed.

Third, it is now widely accepted that the DBS effects are mediated via mechanisms on multiple levels, encompassing the micro-(e.g., local spiking activity), meso-(e.g., local field potentials), and macro-scale (e.g., interregional networks).^22^ In particular, network effects of DBS have received increasing attention in recent years, and it has been postulated that integrating a patient’s connectome into surgical planning could facilitate personalised DBS therapy.^4^ Non-motor outcomes following STN-DBS depend on the location of neurostimulation.^22^ Furthermore, previous studies have associated the stimulation of specific fibre tracts with postoperative outcomes such as depression and impulsivity.^23, 24^ In summary, neuromodulation of subcortical brain regions and connected functional brain networks is associated with non-motor outcomes, whereas cortical atrophy evident in routine MRI scans is not. The clinical implication of our study is that atrophy in the prefrontal cortex should be considered as an MRI biomarker of subpar motor outcome of STN-DBS, whereas cortical atrophy may not indicate worse non-motor outcomes. Compared to these metrics of morphometry based on routine MRI examinations, it is conceivable that more advanced imaging techniques, e.g., markers of cerebral microstructure, such as neurite density, and connectivity measures are more sensitive to map non-motor treatment effects.^25^

### Limitations

First, despite being one of the largest cohorts of its kind, the sample size is relatively small. Nonetheless, it is unlikely that our sample was underpowered because the effect size of postoperative changes of motor and non-motor symptoms was comparable (Cohen’s d: .45, respectively .46) and we observed an association between metrics of morphometry and motor, but not non-motor outcomes. Second, we did not employ scales that specifically measure certain motor and non-motor symptoms, such as the Bain and Findley tremor scale for tremor, the Parkinson’s disease Sleep Scale (PDSS) for sleep, or the Montreal Cognitive Assessment (MoCA) for cognitive symptoms as we were interested in the association between brain morphometry and global motor and non-motor symptom burden. Therefore, we chose the UPDRS-III and the NMSS-T, which represent composite scores for motor and non-motor symptom severity. Furthermore, we wanted to ensure consistency with prior studies employing the UPDRS-III as an outcome parameter. Third, the exact scanning parameters (e.g., repetition-time (TR) and echo-time (TE)) differed slightly across the sample. In a previous study, however, our group demonstrated that results remained consistent even after integrating scanning parameters into the regression analysis.^26^

## Conclusion

Our study supports the importance of intact superior frontal cortex integrity as a predictor for motor outcomes in PD patients undergoing STN-DBS. Despite several advantages, including being based on scans implemented as a standard preoperative procedure, the short scanning time, and established pipelines in analysing and interpreting findings, our results indicate that the use of VBM as a measure to inform the patient selection and preoperative counselling is limited to motor effects and does not extend to non-motor effects of STN-DBS.

## Methods

### Participants

Patients were enrolled in this prospective, observational, ongoing study upon written informed consent in a single centre (University Hospital Cologne). Clinical diagnosis of PD was based on the UK Brain Bank Criteria, and indication for DBS surgery was established according to international guidelines.^27, 28^ Exclusion criteria comprised pathological MR imaging, clinically relevant cognitive impairments, and impaired visual and auditory function. The study was carried out following the Declaration of Helsinki and approved by the University of Cologne ethics committee (study no.: 12-145; German Clinical Trials Register: DRKS00006735).

### Clinical Assessment

Clinical assessments were conducted at the preoperative baseline in the ON-medication state (MedON) and six months after DBS surgery in the ON-medication/ON-stimulation state (MedON/StimON). Standardised case report forms were used to collect demographic and clinical data on both study visits, including a comprehensive neuropsychological assessment which comprised the following scales:

1. The Non-motor Symptoms Scale (NMSS) is a scale evaluated by clinicians that consists of 30 items, which assess nine domains of non-motor symptoms, including (1) cardiovascular, (2) sleep/fatigue, (3) mood/apathy, (4) perceptual problems/hallucinations, (5) attention/memory, (6) gastrointestinal tract, (7) urinary, (8) sexual function, and (9) miscellaneous. The miscellaneous category includes questions regarding pain, the ability to smell/taste, weight change, and excessive sweating. The NMSS has been frequently used in DBS studies for PD.^29–31^ The score on this scale ranges from 0, indicating no impairment, to 360, indicating maximum impairment, while the symptoms are evaluated over the past four weeks.^20^
2. The PD Questionnaire (PDQ)-8 is a self-reported short form of the PDQ-39 that assesses eight dimensions of quality of life (QoL) in patients with PD. The PDQ-8 has been frequently used in DBS studies for PD.^32–34^ The scale is reported as a summary index (SI) and ranges from 0, indicating no impairment, to 100, indicating maximum impairment. ^2, 35, 36^
3. The Unified Parkinson’s Disease Rating Scale part III (UPDRS-III) is a clinician rated scale evaluating motor symptom severity. The UPDRS-III ranges from 0 (no motor impairment) to 108 (maximum motor impairment).{Fahn, 1987 #602}
4. The Scales for Outcomes in PD – Motor Function (SCOPA-M) is a scale evaluated by clinicians that assesses different dimensions of function in PD patients, including activities of daily living and motor complications. The subscale scores range from 0 (no impairment) to 21 for activities of daily living and 12 for motor complications.^37^

The levodopa equivalent daily dose (LEDD) was calculated based on the method described by Jost et al.^38^ Demographic and clinical characteristics are outlined in Table 1.

### MRI Data Acquisition

MRI acquisitions were performed on a 3 T MRI system (Ingenia 3.0T, Philips Healthcare) in a single centre (Cologne). Each PD patient in the MedON underwent a 3D T1-weighted Magnetization Prepared -RApid Gradient Echo sequence (MPRAGE) at baseline.

At the time of image acquisition, images were investigated to be free of motion or ghosting and high frequency or wrap-around artefacts.

### Image Processing

Voxel-based morphometry was performed within the Computational Anatomy Toolbox (CAT) analysis suite (CAT12.8.2, University Hospital Jena, Jena, Germany)^39^ implemented in Statistical Parametric Mapping 12 (SPM12, Wellcome Department of Cognitive Neurology, London, United Kingdom). All steps were conducted in MATLAB R2022a (The MathWorks Inc., Natick, MA, USA), as reported previously by Jergas et al.^26^ In short, processing included spatial registration to a template brain, segmentation into cortical grey matter, white matter, and cerebrospinal fluid, calculation of total intracranial volume (TIV), and empirical quality control (QC) using default parameters. QC was performed within the QC framework of CAT12 with scans not rating lower than B-. Finally, data smoothing was performed using an 8 mm full-width half-maximum isotropic Gaussian kernel.

### Statistical analysis

Statistical analysis of clinical outcomes was performed using MATLAB R2018b. We employed the Shapiro-Wilk test to assess the assumption of normality. Subsequently, Wilcoxon signed-rank- or t-tests, when parametric test criteria were fulfilled, were employed to analyse changes between baseline and 6-month follow-up. The Benjamini-Hochberg method was used to control the false discovery rate and effect sizes were calculated according to Cohen.^40, 41^ Reported p-values are two-sided and were accepted as significant where p<.05. Statistical voxelwise analysis of image data was performed using SPM12. Here, clinical outcomes were represented as change scores in UPDRS-III, NMSS-T, and NMSS-Domains and calculated according to the following equation:

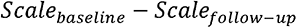

Associations between surrogates of brain morphometry and motor and non-motor outcomes were assessed using a multiple regression analysis with age, sex, and total intracranial volume as covariates. A threshold-free cluster enhancement (TFCE) was applied to correct for multiple comparisons as implemented in the TFCE Toolbox. Results were accepted as significant where family-wise error corrected p<.05.

## Data Availability

All data produced in the present study are available upon reasonable request to the authors

## Acknowledgement

The authors thank the participants for their active engagement in this study.

## Data and Code Availability

The data supporting this study’s findings are available on reasonable request from the corresponding authors (PAL, HSD). The data are not publicly available due to privacy or ethical restrictions. All tools used to analyse MRI data are based on CAT12 (https://neuro-jena.github.io/cat/) and SPM12 (http://www.fil.ion.ucl.ac.uk/spm).

## Contributorship

PAL: study concept and design, data acquisition, data analysis, drafting of the manuscript WS: data acquisition, critical revision of the manuscript

STJ: data acquisition, critical revision of the manuscript

MS: critical revision of the manuscript

JNPS: data acquisition, critical revision of the manuscript

AS: data acquisition, critical revision of the manuscript

AG: data acquisition, critical revision of the manuscript

VVV: data acquisition, surgical intervention, critical revision of the manuscript

GRF: critical revision of the manuscript

JE: critical revision of the manuscript

MK: data acquisition, critical revision of the manuscript

AR: critical revision of the manuscript

AA: critical revision of the manuscript

KA: critical revision of the manuscript

PMM: critical revision of the manuscript

CG: critical revision of the manuscript

KRC: critical revision of the manuscript

LT: study concept and design, critical revision of the manuscript

JCB: study concept and design, data acquisition, data analysis, drafting of the manuscript HSD: study concept and design, data acquisition, data analysis, drafting of the manuscript

## Financial disclosure/Conflicts of Interest

PAL was supported by the SUCCESS-Program of the Philipps-University of Marburg, the ‘Stiftung zur Förderung junger Neurowissenschaftler’, and the Prof. Klaus Thiemann Foundation in the German Society of Neurology.

WS was supported by the Koeln Fortune Program/Faculty of Medicine, University of Cologne and reports no financial relationships with commercial interests.

STJ was funded by the Prof. Klaus Thiemann Foundation and the Brandau-Laibach-Foundation, outside of the submitted work.

MS has received honoraria from Bial, Britannia and Medtronic.

JNPS funded by the Cologne Clinician Scientist Program (CCSP)/Faculty of Medicine/ University of Cologne; and by the German Research Foundation (DFG, FI 773/15-464 1).

AS reports no financial disclosures. AG reports no financial disclosures.

VVV reports compensation for contributions to advisory boards and congresses for Medtronic, Boston Scientific, and Livanova.

GRF serves as an editorial board member of Cortex, Neurological Research and Practice, NeuroImage: Clinical, Zeitschrift für Neuropsychologie, DGNeurologie, and Info Neurologie & Psychiatrie; receives royalties from the publication of the books Funktionelle MRT in Psychiatrie und Neurologie, Neurologische Differentialdiagnose, and SOP Neurologie; receives royalties from the publication of the neuropsychological tests KAS and Köpps; received honoraria for speaking engagements from Deutsche Gesellschaft für Neurologie

(DGN) and Forum für medizinische Fortbildung FomF GmbH.

JE reports no financial disclosures.

MK was funded by the KoelnFortune program of the Medical Faculty of the University of Cologne and reports no financial relationships with commercial interests.

AR received funding from the National Institute for Health and Care Research (NIHR), the Clinical Research Network (CRN) South London, and grant funding from the International Parkinson and Movement Disorders Society (MDS).

AA received compensation for consultancy and speaker-related activities from UCB, Boehringer Ingelheim, Ever Pharma, General Electric, Britannia, AbbVie, Kyowa Kirin, Zambon, Bial, Theravance Biopharma, Jazz Pharmaceuticals, Roche, Medscape. He receives research support from Bial, Lundbeck, Roche, Angelini Pharmaceuticals, Horizon 2020 Grant 825785, Horizon 2020 Grant 101016902, Ministry of Education University and Research (MIUR) Grant ARS01_01081, Cariparo Foundation, Movement Disorders Society for NMS Scale validation. He serves as consultant for Boehringer–Ingelheim for legal cases on pathological gambling.

KA has received honoraria for educational meetings, travel and consultancy from Medtronic, St. Jude Medical and Boston Scientific.

PMM received honoraria from the International Parkinson and Movement Disorder Society (IPMDS) for management of the Clinical Outcome Assessment Program.

CG reports no financial disclosures.

KRC received grants (IIT) from Britannia Pharmaceuticals, AbbVie, UCB, GKC, EU Horizon 2020, Parkinson’s UK, NIHR, Parkinson’s Foundation, Wellcome Trust, Kirby Laing Foundation, MRC; royalties or licenses from Oxford (book), Cambridge publishers (book), MAPI institute (KPPS, PDSS 2); consulting fees, support for attending meetings or travel, and participated on data safety monitoring board or advisory board for AbbVie, UCB, GKC, Bial,

Cynapsus, Lobsor, Stada, Zambon, Profile Pharma, Synovion, Roche, Therevance, Scion, Britannia, Acadia, 4D Pharma, and Medtronic; and served as a committee chair for MDS (unpaid) and EAN (unpaid).

LT received payments as a consultant for Medtronic Inc. and Boston Scientific and received honoraria as a speaker on symposia sponsored by Bial, Zambon Pharma, UCB Schwarz Pharma, Desitin Pharma, Medtronic, Boston Scientific, and Abbott. The institution of LT, not LT personally, received funding by the German Research Foundation, the German Ministry of Education and Research, and Deutsche Parkinson Vereinigung.

JCB is funded by the Else Kroener-Fresenius-Stiftung (2022_EKES.23) and receives funding by the German Research Foundation (Project ID 431549029-C07).

HSD was funded by the EU Joint Programme – Neurodegenerative Disease Research (JPND), the Prof. Klaus Thiemann Foundation in the German Society of Neurology, the Felgenhauer Foundation, the KoelnFortune program of the Medical Faculty of the University of Cologne and has received honoraria by Everpharma, Kyowa Kirin, Bial, Oruen, and Stadapharm.

## Notes

### Competing Interest Statement

The authors have declared no competing interest.

### Funding Statement

PAL was supported by the SUCCESS-Program of the Philipps-University of Marburg, the Stiftung zur Foerderung junger Neurowissenschaftler, and the Prof. Klaus Thiemann Foundation in the German Society of Neurology.
WS was supported by the Koeln Fortune Program/Faculty of Medicine, University of Cologne and reports no financial relationships with commercial interests.
STJ was funded by the Prof. Klaus Thiemann Foundation and the Brandau-Laibach-Foundation, outside of the submitted work.
MS has received honoraria from Bial, Britannia and Medtronic.
JNPS funded by the Cologne Clinician Scientist Program (CCSP)/Faculty of Medicine/ University of Cologne; and by the German Research Foundation (DFG, FI 773/15-464 1).
AS reports no financial disclosures.
AG reports no financial disclosures.
VVV reports compensation for contributions to advisory boards and congresses for Medtronic, Boston Scientific, and Livanova.
GRF serves as an editorial board member of Cortex, Neurological Research and Practice, NeuroImage: Clinical, Zeitschrift fuer Neuropsychologie, DGNeurologie, and Info Neurologie & Psychiatrie; receives royalties from the publication of the books Funktionelle MRT in Psychiatrie und Neurologie, Neurologische Differentialdiagnose, and SOP Neurologie; receives royalties from the publication of the neuropsychological tests KAS and Koepps; received honoraria for speaking engagements from Deutsche Gesellschaft fuer Neurologie (DGN) and Forum fuer medizinische Fortbildung FomF GmbH.
JE reports no financial disclosures.
MK was funded by the KoelnFortune program of the Medical Faculty of the University of Cologne and reports no financial relationships with commercial interests.
AR received funding from the National Institute for Health and Care Research (NIHR), the Clinical Research Network (CRN) South London, and grant funding from the International Parkinson and Movement Disorders Society (MDS).
AA received compensation for consultancy and speaker-related activities from UCB, Boehringer Ingelheim, Ever Pharma, General Electric, Britannia, AbbVie, Kyowa Kirin, Zambon, Bial, Theravance Biopharma, Jazz Pharmaceuticals, Roche, Medscape. He receives research support from Bial, Lundbeck, Roche, Angelini Pharmaceuticals, Horizon 2020 Grant 825785, Horizon 2020 Grant 101016902, Ministry of Education University and Research (MIUR) Grant ARS01 01081, Cariparo Foundation, Movement Disorders Society for NMS Scale validation. He serves as consultant for Boehringer-Ingelheim for legal cases on pathological gambling.
KA has received honoraria for educational meetings, travel and consultancy from Medtronic, St. Jude Medical and Boston Scientific.
PMM received honoraria from the International Parkinson and Movement Disorder Society (IPMDS) for management of the Clinical Outcome Assessment Program.
CG reports no financial disclosures.
KRC received grants (IIT) from Britannia Pharmaceuticals, AbbVie, UCB, GKC, EU Horizon 2020, Parkinson's UK, NIHR, Parkinson's Foundation, Wellcome Trust, Kirby Laing Foundation, MRC; royalties or licenses from Oxford (book), Cambridge publishers (book), MAPI institute (KPPS, PDSS 2); consulting fees, support for attending meetings or travel, and participated on data safety monitoring board or advisory board for AbbVie, UCB, GKC, Bial,
Cynapsus, Lobsor, Stada, Zambon, Profile Pharma, Synovion, Roche, Therevance, Scion, Britannia, Acadia, 4D Pharma, and Medtronic; and served as a committee chair for MDS (unpaid) and EAN (unpaid).
LT received payments as a consultant for Medtronic Inc. and Boston Scientific and received honoraria as a speaker on symposia sponsored by Bial, Zambon Pharma, UCB Schwarz Pharma, Desitin Pharma, Medtronic, Boston Scientific, and Abbott. The institution of LT, not LT personally, received funding by the German Research Foundation, the German Ministry of Education and Research, and Deutsche Parkinson Vereinigung.
JCB is funded by the Else Kroener-Fresenius-Stiftung (2022 EKES.23) and receives funding by the German Research Foundation (Project ID 431549029-C07).
HSD was funded by the EU Joint Programme Neurodegenerative Disease Research (JPND), the Prof. Klaus Thiemann Foundation in the German Society of Neurology, the Felgenhauer Foundation, the KoelnFortune program of the Medical Faculty of the University of Cologne and has received honoraria by Everpharma, Kyowa Kirin, Bial, Oruen, and Stadapharm.

### Author Declarations

The ethics committee of the University of Cologne gave approval for this work.

